# Neuromagnetic speech discrimination responses are associated with reading-related skills in dyslexic and typical readers

**DOI:** 10.1101/2019.12.29.19016113

**Authors:** A. Thiede, L. Parkkonen, P. Virtala, M. Laasonen, J.P. Mäkelä, T. Kujala

## Abstract

Dyslexia is thought to result from poor phonological processing. We investigated neuromagnetic speech discrimination in dyslexic and typical readers with mismatch fields (MMF) and determined the associations between MMFs and reading-related skills. We expected weak and atypically lateralized MMFs in dyslexic readers, and associations between reading-related skills and MMF strength. MMFs were recorded to a repeating pseudoword /*ta-ta*/ with occasional changes in vowel identity, duration, or syllable frequency from 43 adults, 21 with confirmed dyslexia. Speech-sound changes elicited MMFs in bilateral auditory cortices, with no group differences in source strengths. MMFs to vowel identity and duration changes were left-lateralized. Better verbal working memory was associated with stronger left-hemispheric MMFs across groups, suggesting the relevance of verbal working memory for speech processing. Better technical reading was associated with stronger right-hemispheric MMFs in dyslexic readers, suggesting a right-hemispheric compensatory mechanism for language processing. In conclusion, contrary to prior work, our results did not support deficient speech discrimination in dyslexia. However, in line with previous studies, we observed left-lateralized MMFs to vowel identity and duration changes, and associations of MMFs with reading-related skills, highlighting the connection between neural speech processing and reading and promoting the use of MMFs in investigating reading-related brain processes.

**Highlights:**

- Speech-sound changes elicit comparable mismatch fields in dyslexics and controls.
- Mismatch fields (MMFs) to vowel identity and duration changes are left-lateralized.
- Stronger left MMFs are associated with better verbal working memory across groups.
- Stronger right MMFs are associated with better technical reading in dyslexics.
- Low-level neural speech discrimination is associated with reading-related skills.

## 1 Introduction

Developmental dyslexia has been associated with several structural and functional brain abnormalities and significant difficulties in phonological processing (Laasonen, Lehtinen, Leppämäki, Tani, & Hokkanen, 2010; Ramus, 2001; Ramus, Altarelli, Jednoróg, Zhao, & Scotto di Covella, 2018), which may result from poor phonological representations or their accessibility (Ramus, 2001; Ramus, Marshall, Rosen, & van der Lely, 2013). Also working memory, especially verbal short-term memory is impaired in at least some dyslexic readers (Banai & Ahissar, 2004; Laasonen, Leppämäki, Tani, & Hokkanen, 2009).

Acoustic–phonological processes pertinent for speech functions can be investigated with mismatch negativity (MMN) with electroencephalography (EEG) or magnetoencephalography (MEG; Näätänen, 2001). MMN is an event-related component elicited by rare changes in a stream of repeating sounds (Näätänen, Gaillard, & Mäntysalo, 1978), reflecting neural sound discrimination (Kujala & Näätänen, 2010). The primary MMN generators are located in bilateral temporal cortices (e.g., Alho, 1995), with the left generator having a stronger contribution to speech processing than the right one (e.g., Näätänen et al., 1997; Shtyrov, Kujala, Palva, Ilmoniemi, & Näätänen, 2000).

Larger MMN amplitudes to changes in speech sounds have been associated with better phoneme processing skills in typically developing prereaders (Linnavalli, Putkinen, Huotilainen, & Tervaniemi, 2017) and better nonword scores in children with auditory processing disorder (Sharma et al., 2006). MMNs can also predict future development, as shown by MMNs recorded to speech sounds in kindergarten (Maurer et al., 2009) and in infancy (Van Zuijen, Plakas, Maassen, Maurits, & Van der Leij, 2013) that are associated with language and reading outcomes at school. Furthermore, grapheme-phoneme mapping intervention of preschoolers at risk of dyslexia increased the MMN to vowel changes in syllables concurrently with the improvement of pre-reading skills (Lovio, Halttunen, Lyytinen, Näätänen, & Kujala, 2012). These results suggest that there is an association between MMN and language and reading skills. In line with this, MMNs to speech-sound and non-speech sound changes were shown to be diminished and delayed in children and adults with dyslexia (for reviews, see Hämäläinen, Salminen, & Leppänen, 2013; Kujala & Näätänen, 2001; Schulte-Körne & Bruder, 2010), and even in infants and children having a familial risk of dyslexia (Benasich et al., 2006; Leppänen et al., 2002; Lovio, Näätänen, & Kujala, 2010; Thiede et al., 2019; van Leeuwen et al., 2008; for a review, see Ozernov-Palchik & Gaab, 2016). Not all studies, however, were able to consistently show such results (Corbera, Escera, & Artigas, 2006; Hämäläinen, Leppänen, Guttorm, & Lyytinen, 2008; Meng et al., 2005; Schulte-Körne, Deimel, Bartling, & Remschmidt, 1998), suggesting that some types of changes or change magnitudes might not be sensitive enough to probe the neural auditory/speech deficit in dyslexia.

Also abnormal MMN lateralization to tone changes in dyslexia has been found (Kujala, Belitz, Tervaniemi, & Näätänen, 2003; Kujala et al., 2000; Renvall & Hari, 2003; Sebastian & Yasin, 2008; see, however, Kujala, Lovio, Lepistö, Laasonen, & Näätänen, 2006; Schulte-Körne, Deimel, Bartling, & Remschmidt, 1999, 2001; Sharma et al., 2006). Only few studies have investigated lateralization of speech-elicited MMNs in dyslexia or at risk of dyslexia, the results being mixed. Left-lateralized MMN to phoneme changes in kindergarten was found to predict good and right-lateralized MMN poor reading skills at school (Maurer, Bucher, Brem, & Brandeis, 2003). Some studies report no differences in the lateralization of the speech-elicited MMN between dyslexic and control groups (Schulte-Körne et al., 2001; Sebastian & Yasin, 2008).

Very few MMN studies on dyslexia have so far used spatially accurate methods to determine response strengths or lateralization. Renvall and Hari (2003) reported weaker left-hemispheric mismatch fields (MMFs, used here) or magnetic mismatch negativity (MMNm) to tone frequency changes in adult dyslexic compared to non-dyslexic readers. To our knowledge, the only previous study comparing MMFs in dyslexic and control participants to speech-sound changes (/*ba*/ vs. /*da*/) failed to find group differences (Paul, Bott, Heim, Wienbruch, & Elbert, 2006). The present study addresses this apparent niche in dyslexia research by utilizing MEG with improved source localization applying individual head models from anatomical MRIs.

We investigated neural speech discrimination in dyslexia and its association with reading-related skills by recording MMFs to several relevant speech-sound changes (vowel, vowel duration, syllable frequency) in a phonotactically legal pseudoword. Furthermore, we used an extensive neuropsychological test battery tapping different cognitive domains including reading, phonological processing, intelligence quotient (IQ), and working memory. We expected to find diminished and/or delayed MMF source amplitudes in dyslexic participants. Based on previous studies on lateralization, no clear prediction could be made on group laterality differences. Furthermore, better outcomes in reading-related neuropsychological tests were expected to correlate with stronger MMFs. To our knowledge, this is the first study determining neural sources of phonetic discrimination in adults with dyslexia.

## 2 Material and Methods

### 2.1 Participants

Forty-three healthy Finnish participants (21 dyslexics, 22 controls) aged 19–45 years without history of neurological diseases participated in the study. The inclusion criteria for the dyslexic group were a diagnostic dyslexia statement (from a psychologist, special education teacher or similar), or a history of reading difficulties in childhood (see 2.2) combined with below-norm performance in either speed or accuracy (below one standard deviation from age-matched standardized control data, see Laasonen et al., 2010) in two or more reading subtests (word list reading, pseudoword list reading, text reading, Nevala, Kairaluoma, Ahonen, Aro, & Holopainen, 2006). Inclusion criteria for control participants were no report of dyslexia or co-occurring language disorders confirmed by within-norm performance in speed and accuracy in at least two reading subtests. General exclusion criteria were attention deficit disorders (see 2.2), performance IQ below 80, metal in the body, and an individualized school curriculum. Participants gave their written informed consent, and all procedures employed conformed to the Declaration of Helsinki. The Coordinating Ethics Committee (Hospital District of Helsinki and Uusimaa) approved the study protocol. The study has been pre-registered in ClinicalTrials.org (ID <u>NCT02622360</u>).

### 2.2 Questionnaires and neuropsychological test battery

All participants filled questionnaires before brain imaging measurements including Finnish versions of The Adult Reading History Questionnaire (ARHQ; Laasonen et al., 2014) and The Adult ADHD Self-Report Scale (ASRS; Kessler et al., 2005), paired with interviews. The neuropsychological tests included domains of phonological processing (span length in Nonword span, Laasonen, 2002; accuracy in Pig Latin, Nevala et al., 2006; speed in the second trial of Rapid Alternate Stimulus Naming RAN, Wolf, 1986), technical reading (speed and accuracy in word list reading and pseudoword list reading, Nevala et al., 2006), intelligence (verbal, performance, and full IQ calculated from subtests Similarities and Vocabulary, Block design and Matrix reasoning, and across all four, respectively, in the Wechsler Adult Intelligence Scale III, Wechsler, 2005) and working memory (visual working memory [Visual Series] and verbal working memory [Letter-Number Series subtest] of the Wechsler Memory Scale III, WMS-III, Wechsler, 2008). Composite scores were computed for phonological processing and technical reading by converting the raw scores of all subtests to z-scores and averaging them, and for working memory functions by following the procedure advised in WMS-III.

The control group had higher education than the dyslexia group and outperformed the dyslexia group in technical reading, phonological processing, working memory as well as in verbal and performance IQ (Table 1, Figure S 1). As dyslexic readers are known to underperform in verbal, but not necessarily in performance IQ (Laasonen et al., 2009), the full IQ was taken into account in the correlation analysis as a control variable, and analyses including group comparisons were repeated with groups matched for performance IQ (see 2.6).

**Table 1.** Demographic and neuropsychological characterization of the study groups.

| Variable | CON (N=22) | DYS (N=21) | comparison |  |  |
| --- | --- | --- | --- | --- | --- |
|  |  |  | test statistic | p | 95% confidence interval |
| **demographic** |  |  |  |  |  |
| gender [f/m] | 12/10 | 12/9 | $\chi^2(1) = 0$ | 1 | |
| age [years] | 29.9 (5.9) | 31.0 (8.6) | $t(35) = -0.48$ | .633 | [-5.68 3.50] |
| education [years] | 17.0 (2.5) | 14.7 (2.5) | $t(40) = 3.06$ | **.004 | [0.80 3.93] |
| music education<sup>#</sup> [years] | 0.25 (5.5) | 0.0 (2.0) | $W = 283.5$ | .152 | |
| **neuropsychological** |  |  |  |  |  |
| full IQ | 117.0 (7.0) | 104.0 (9.4) | $t(37) = 5.18$ | ***$8.0 \cdot 10^{-6}$ | [8.00 18.26] |
| verbal IQ<sup>#</sup> | 115.0 (10.0) | 100.0 (22.0) | $W = 390.5$ | ***$1.0 \cdot 10^{-4}$ | |
| performance IQ | 120.0 (10.0) | 110.0 (12.3) | $t(39) = 3.13$ | **.003 | [3.77 17.61] |
| phonological processing | 0.49 (0.42) | -0.33 (0.60) | $t(36) = 5.18$ | ***$8.9 \cdot 10^{-6}$ | [0.50 1.14] |
| technical reading<sup>#</sup> | 0.61 (0.17) | -0.34 (0.82) | $W = 461$ | ***$3.8 \cdot 10^{-12}$ | |
| working memory | 24.3 (4.8) | 19.8 (5.0) | $t(41) = 3.01$ | **.004 | [1.49 7.53] |

Notes. Mean and standard deviation (in brackets) for normally-distributed variables (Shapiro-Wilk test). Group comparison with independent-samples t-test. Median and interquartile range (in brackets) for non-normally-distributed variables, indicated by the hash sign<sup>#</sup>. Group comparison with Wilcoxon Rank Sum W-test. Test statistics, degrees of freedom (in brackets), significance levels *p*, and 95% confidence intervals are reported. \**p* < .05, \*\**p* < .01, \*\*\**p* < .001. CON – control group, DYS – dyslexic group.

### 2.3 Stimuli

The repetitive “standard” stimulus was a Finnish 300-ms-long pseudoword /*ta-ta*/ (75.3%) with the stress on the first syllable. Occasional “deviant” stimuli (8.3% of each type) included a change in vowel duration (lengthening of second /*a*/ from 71 to 158 ms), vowel identity (adding /*o*/ to the second syllable from a natural recording of /*ta-to*/, pitch-controlled), or syllable frequency (shifting the F0 of the second syllable from 175 to 225 Hz) in the second syllable (edited with Adobe Audition CS6, 5.0, Build 708 and Praat 5.4.01). Stimuli were presented pseudo-randomly (at least one standard always following a deviant) in two ≈12.6 min recording blocks, each containing 946 stimuli and starting with five standard stimuli. The stimulus-onset asynchrony (SOA) was 800±50 ms (randomly alternating between 750, 760, 770, …, 840, 850 ms).

The stimuli were presented with Presentation Software (Neurobehavioural Systems Ltd., Berkeley, CA, USA) binaurally via plastic tubes and silicon earphones at a comfortable level (≈70–80 dB SPL). During stimulation, participants were sitting, instructed to keep the head still and to attend to a silenced movie projected (Panasonic PT-D7500E; Panasonic, Kadoma, Osaka, Japan) to a back-projection screen (MEGIN Oy, Helsinki, Finland) located 150 cm from the participant’s head.

### 2.4 MEG/EEG and MRI procedure

MEG/EEG was recorded using a 306-channel Elekta Neuromag TRIUX (MEGIN Oy, Helsinki, Finland) whole-head MEG system (sampling rate 1 kHz and pass-band 0.03–330 Hz) in a magnetically shielded room (Euroshield/ETS Lindgren Oy, Eura, Finland) in BioMag Laboratory in Helsinki University Central Hospital (duration 2–3 h). Prior to the measurement, the positions of five head position indicator (HPI) coils and additional head surface points (EEG electrodes) were determined in relation to the nasion and both preauricular points with an Isotrak 3D-digitizer (Polhemus Inc., Colchester, USA). The head position with respect to the MEG sensor array was continuously monitored. Vertical and horizontal electro-oculograms (EOG) were recorded.

The anatomical T1-weighted images (MPRAGE) were acquired on a 3T MAGNETOM Skyra whole-body MRI scanner (Siemens Healthcare, Erlangen, Germany) with a 32-channel head coil at AMI centre of Aalto Neuroimaging (duration 30 min), Aalto University (176 slices, slice thickness 1 mm, voxel size 1 mm x 1 mm x 1 mm, field of view 256 mm x 256 mm). The images were checked for incidental findings by a physician.

### 2.5 Data preprocessing

The following processing steps were executed in MNE-Python software package v0.17.dev0 (Gramfort et al., 2014), unless indicated otherwise; the code is available at https://github.com/athiede13/neural_sources. Temporal signal space separation (tSSS; Taulu & Simola, 2006) with head movement compensation and interpolation of previously marked bad channels was performed with Maxfilter software (version 2.2.15; MEGIN Oy, Helsinki, Finland). Ocular and cardiac artifacts were removed by signal space projection (SSP; Tesche et al., 1995). The data were filtered to 0.5–30 Hz with a finite impulse response filter, and epochs were extracted 100 ms before and 840 ms after stimulus onset for all stimulus types for all channels and each participant and recording. Epochs with signal excursion exceeding 4 pT in magnetometers, 4 pT/cm in gradiometers and 250 µV in EOG channels were excluded from analysis. For the standard stimuli, on average 679 epochs (range 667–681), and for deviants, on average 144 epochs (range 138–147) per participant were included in the analysis. Deviant-minus-standard subtraction curves (MMFs) were calculated for each deviant type with equal weights.

The anatomical MRI of each participant was preprocessed with Freesurfer software (http://surfer.nmr.mgh.harvard.edu/) version 5.3 and 6.0 following the standard procedure (Fischl, 2012). Manual editing of pial surface and white matter control points (64% and 18% of cases, respectively) ensured a correct segmentation of the cortex.

### 2.6 Source modeling

After MRI–MEG coregistration (*mne coreg*), the generators of individual MMFs were estimated in a cortically-constrained source space; the MEG forward solution was calculated for 4098 source points per hemisphere. The minimum-norm source estimate (MNE) was computed for the MMFs using depth-weighting (0.8), fixed-orientation constraint, and the 100-ms-pre-stimulus-baseline regularized noise covariance of pooled deviant and standard waveforms of each participant. The MNE source estimates were morphed to Freesurfer’s average subject (*fsaverage*) cortical space and averaged for each group.

Auditory cortices (lateral sections of superior temporal gyrus and sulcus of the Destrieux Atlas, aparc.a2009s in Freesurfer, Destrieux, Fischl, Dale, & Halgren, 2010) were *a priori* chosen as regions of interest (ROI) based on previous research (Alho, 1995; Renvall & Hari, 2003). As this region is considerably larger than the presumed region generating MMF, the average of the dipole moments within that ROI is not representative of the activity of interest. Therefore, data-driven functional ROIs were created based on the MNE source estimate of each participant for each MMF at the individual peak time within the MMF time window (300– 400 ms after stimulus onset). Specifically, the functional ROIs were created by taking the top 60% of individual peak source estimates within the anatomical *a priori* -defined ROI and then combined into one for the most consistent individual source points (in more than 45% of all functional ROIs) to represent the area with the most consistent activity across participants. The source time course at the functional ROI were extracted with source signs flipped depending on the source orientation (*pca_flip*) to avoid cancellation.

Time windows for determining the maximal MMF source amplitudes were selected around the peaks of the group averages across groups and deviant types, resulting in a 100-ms wide window for the first peak (300–400 ms) and a 200-ms wide window for the second peak (450–650 ms). Individual maximum amplitudes and the corresponding latencies were extracted within the determined time windows and ROI for statistical analysis in both hemispheres.

### 2.7 Statistical analysis

SPSS version 25.0.0.1 (IBM, Armonk, New York, USA), R (R Core Team, 2018) and RStudio version 1.1.453 (RStudio Team, 2016) were used for statistical analyses. Group and laterality effects and their interactions on MMF source amplitudes and latencies were tested for significance with one-sample *t*-tests and analyzed with two-way repeated-measures analysis of variance (ANOVA) with group (control, dyslexic) as between-subjects factor and laterality (left, right) as within-subjects factor, separately for the three deviants and the two time windows.

Partial Pearson correlations were computed between MMF source amplitudes and neuropsychological test scores, controlling for the effect of full IQ. In order to reduce the amount of tests, correlations were analyzed in three steps both separately for the two groups and across groups: (1) MMF source amplitudes at two hemispheres averaged across all deviants with the three neuropsychological composite scores, if significant, then (2) separately for MMFs to each deviant, if significant, then (3) separately for each subcomponent of the neuropsychological composite scores. Despite a potential risk of circular inference (Kriegeskorte, Simmons, Bellgowan, & Baker, 2009), this stepwise procedure was chosen to output the most meaningful associations from a neuroscientific and neuropsychological perspective. A correction for multiple comparisons is addressed here unlike in many brain-behavior correlational studies (Rousselet & Pernet, 2012).

Lateralization indices (LI) for the MMF source amplitudes were computed as LI = (left –right)/(left + right). An LI ≤ –0.20 was considered as right-dominance, LI ≥ 0.20 as left-dominance, and LI between those values as bilateral (Han & Dimitrijevic, 2015). LI values were compared between groups with independent-samples *t*-tests.

To ensure that the IQ difference would not explain the results, statistical analyses comparing groups were repeated for a sample in which the groups were matched for performance IQ (*N* = 37, 19 in control group, 18 in dyslexic group). The profile of the matched groups did not otherwise differ from the original sample.

## 3 Results

MEG source estimates in both groups (Figure 1) revealed that all speech-sound changes elicited MMFs in the auditory cortices. Peak activations were located in the left middle temporal cortex (BA21 and BA22) and the right superior temporal cortex (BA41 and BA22). The source waveform indicated two responses, referred to as MMF and late MMF. Both responses were statistically significantly larger than zero in both groups and for all deviants (for all *p* < .002).

**Figure 1.**
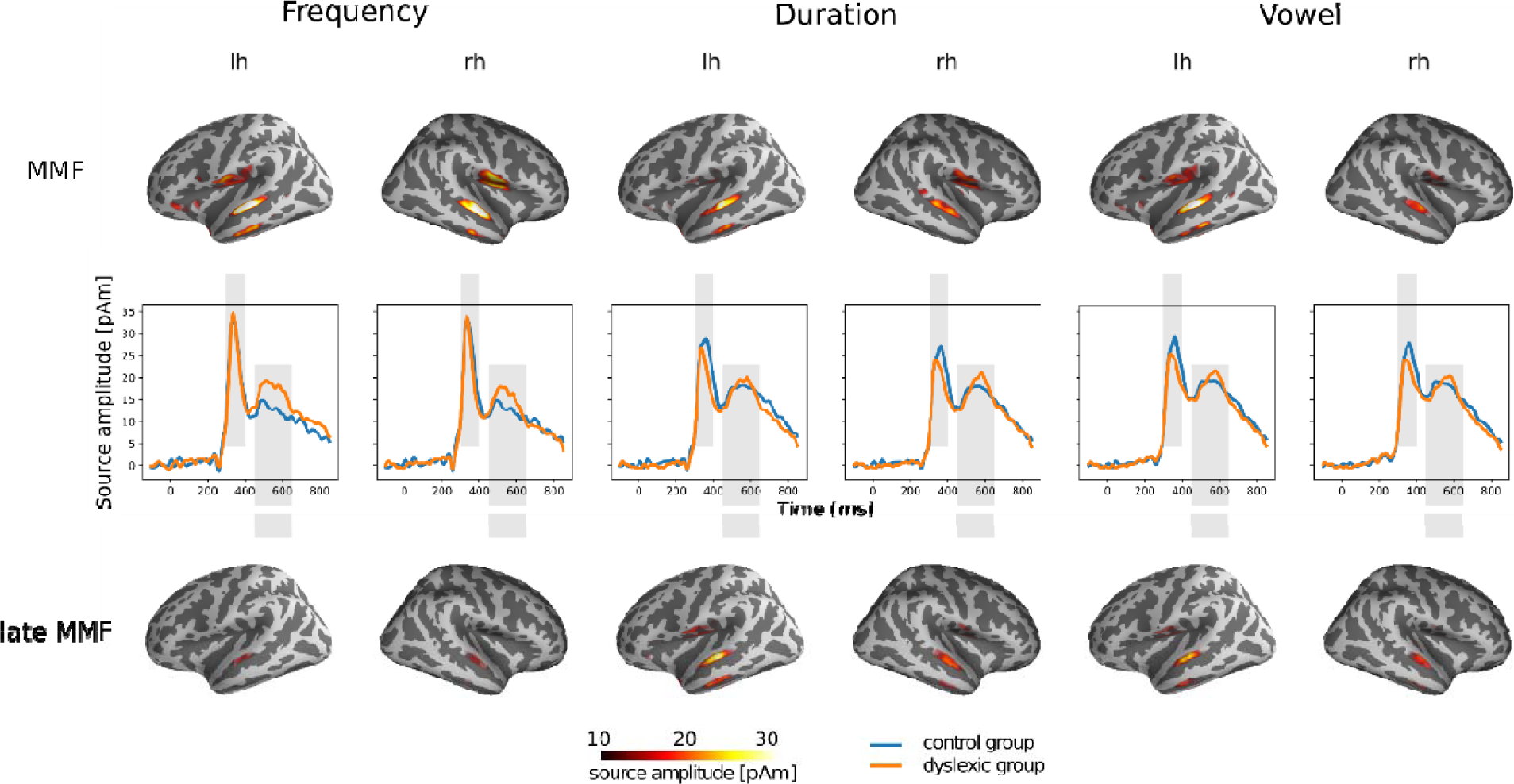
Neural sources of MMF and late MMF to frequency, duration, and vowel changes as MNE’s on lateral brain surfaces (top and bottom panel), and extracted ROI time courses (middle panel). Grey shaded areas in the middle panel depict the time windows of interest (first – MMF, second – late MMF).

A significant laterality main effect was found for the MMF source amplitudes to the vowel deviant, indicating larger left-than right-hemispheric MMFs (*F*(1,41) = 19.12, *p* = 8.2*10^−5^, *p*_adj_ = 4.9*10^−4^, mean difference MD = 14 pAm). For the late MMF source amplitudes, significant laterality main effects were found for duration (*F*(1,41) = 10.05, *p* = .003, *p*_adj_ = .018, MD = 9 pAm) and vowel deviants (*F*(1,41) = 11.62, *p* = .001, *p*_adj_ = .006, MD = 11 pAm), indicating larger late MMFs in the left than right hemispheres. A significant laterality main effect for the late duration MMF latency was found, with a faster right-than left-hemispheric response (*F*(1,41) = 4.12, *p* = .049), however, it was not significant after Bonferroni-correction, (*p*_adj_ = .294, MD = 16 ms). Full ANOVA statistics are reported in Table 2.

**Table 2.** Significant ANOVA results (N = 43)

| comp.<br>dev. | ANOVA |  |  |  |  |  | Post-hoc |  |  |
| --- | --- | --- | --- | --- | --- | --- | --- | --- | --- |
|  | model | effect | F(1,41) | p | p<sub>corr</sub> | η<sub>p</sub><sup>2</sup> | effect | MD | stat |
| **MMF** |  |  |  |  |  |  |  |  |  |
| fre | laterality * amplitude * group | laterality | 3.95 | .054 | .324 | .09 | left > right | 7 pAm |  |
| dur | laterality * amplitude * group | group | 3.61 | .064 | .384 | .08 | con > dys | 7 pAm |  |
| dur | laterality * latency * group | laterality | 2.99 | .091 | .546 | .07 | left > right | 8 ms |  |
| vow | laterality * amplitude * group | laterality | 19.12 | 8.2*10<sup>-5</sup> | ***4.9*10<sup>-4</sup> | .32 | left > right | 14 pAm |  |
| **late MMF** |  |  |  |  |  |  |  |  |  |
| fre | laterality * amplitude * group | laterality | 3.00 | .091 | .546 | .07 | left > right | 6 pAm |  |
| fre | laterality * latency * group | laterality *<br>group | 4.03 | .051 | .306 | .09 | right: con < dys | -27 ms | t(41) = -1.83,<br>p = .074 |
|  |  |  |  |  |  |  | con: left > right | 34 ms | t(21) = 1.92,<br>p = .069 |
| dur | laterality * amplitude * group | laterality | 10.05 | .003 | *.018 | .20 | left > right | 9 pAm |  |
| dur | laterality * latency * group | laterality | 4.12 | .049 | .294 | .09 | left > right | 16 ms |  |
| vow | laterality * amplitude * group | laterality | 11.62 | .001 | ** .006 | .22 | left > right | 11 pAm |  |

Notes. Significant ( $p < .05$ ) and marginally significant ( $p < .1$ ) uncorrected $p$ -values are reported. Bonferroni-corrected significance levels are
indicated with asterisks \* $p < .05$ , \*\* $p < .01$ , \*\*\* $p < .001$ . comp. – component, dev. – deviant, dur – duration, vow – vowel, fre – frequency, MD –
mean difference, stat – full statistics, con – control group, dys – dyslexic group.

Lateralization indices (LIs) for MMF and late MMF did not significantly differ between the groups (*p* > .40). The LIs indicate that the MMF was left-lateralized for the frequency deviant in the control (con) group (mean LI 0.21, SD 0.63) and bilateral in the dyslexic (dys) group (0.08, 0.36). Both groups had bilateral MMFs for the duration deviant (con: 0.05, 0.34; dys: -0.03, 0.33) and left-lateralized MMF for the vowel deviant (con: 0.20, 0.43; dys: 0.23, 0.38). LIs for the late MMF were consistent across groups. It was right-lateralized for the frequency deviant (con: -0.53, 9.02; dys: -1.83, 7.69) and left-lateralized for duration (con: 1.45, 6.12; dys: 0.29, 1.69) and vowel (con: 0.52, 1.63; dys: 0.88, 2.38) deviants.

Source amplitudes of MMF and late MMF correlated with neuropsychological test scores (Figure 2, for complete statistics, see Table S 2). Larger MMF source amplitudes in the left hemisphere were weakly correlated with better working memory skills across all deviants and across both groups (*r* = 0.24, *p*_adj_ = .033). Separate correlation tests for the deviants revealed a moderate correlation across groups for the left duration MMF with working memory skills (*r* = 0.38, *p*_adj_ = .036). Separate working memory subtest analysis (visual and verbal) only showed a significant moderate correlation between the left duration MMF and the verbal working memory component (*r* = 0.45, *p*_adj_ = .006). Within the control group, left MMFs across all deviants correlated moderately with working memory skills (*r* = 0.36, *p*_adj_ = .020). In separate tests for the deviants, however, none remained significant. Within the dyslexic group, larger right MMFs (*r* = 0.35, *p*_adj_ = .039) and right late MMFs (*r* = 0.41, *p*_adj_ = .008) across all deviants correlated moderately strongly with better technical reading skills. In separate tests for the deviants, however, none remained significant.

**Figure 2.**
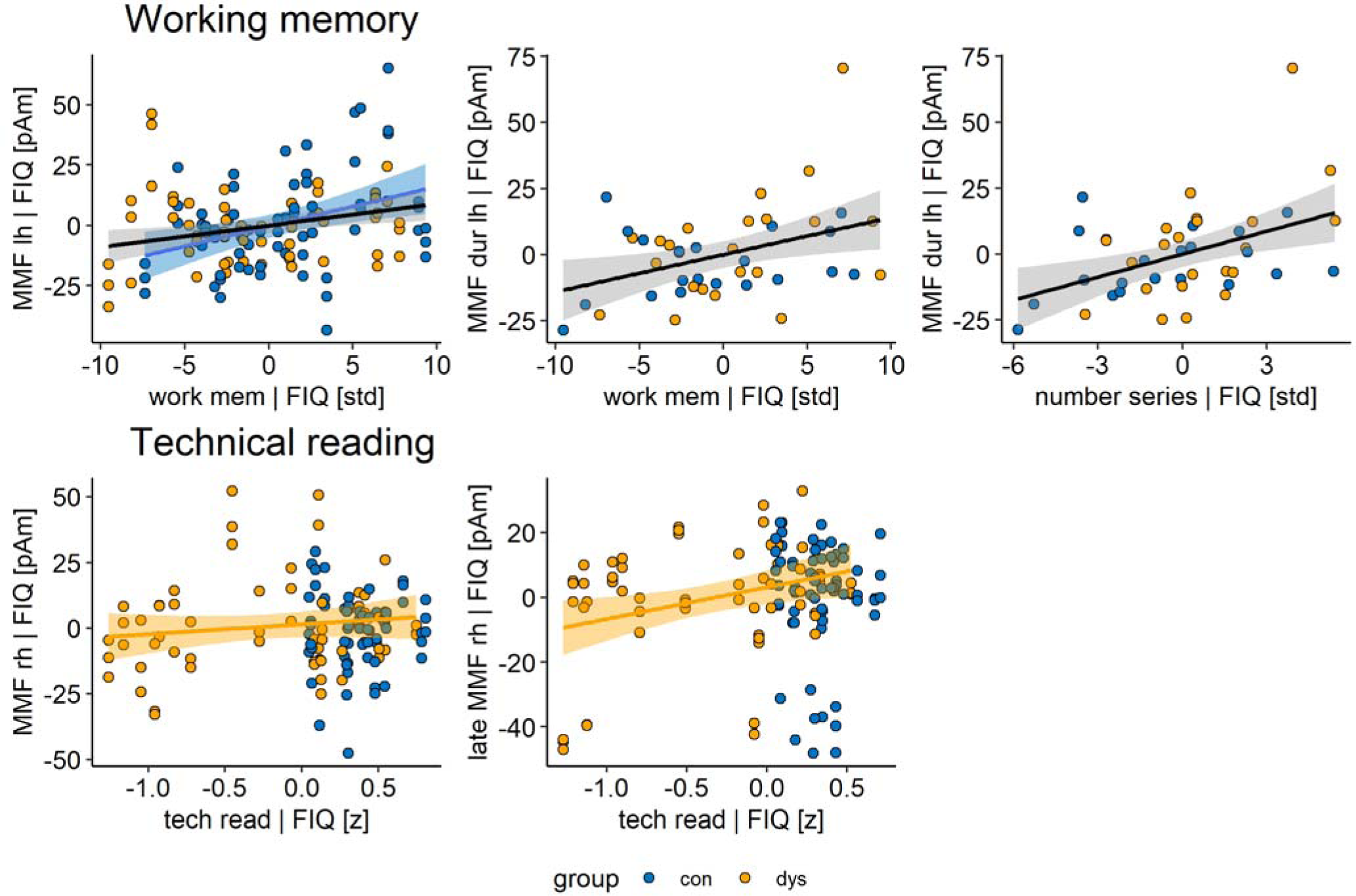
Significant partial Pearson correlations after Bonferroni correction of MMF and late MMF source amplitudes with working memory and technical reading skills after regressing out FIQ. Scatter plots with linear regression lines (black – across both groups, blue – control group, yellow – dyslexic group) are shown for both all deviants pooled together (upper left and both bottom panels) and separately for one deviant (upper middle and right panels). Number series is one subcomponent of the working memory composite score (upper right panel). One outlier (dyslexic) was removed, because of the technical reading score being below three interquartile ranges. work mem – working memory, tech read – technical reading, dur - duration, lh – left hemisphere amplitude, rh – right hemisphere amplitude

The reported ANOVA results remained similar when repeating the analyses for performance-IQ-matched subsamples of control and dyslexic groups (Table S 3), and LIs did not differ significantly between groups (*p* > .40).

## 4 Discussion

Our goals were to determine whether neural speech-sound discrimination, reflecting phonetic representations, is deficient or abnormally lateralized in adult dyslexic readers, and whether the neural responses correlate with reading-related skills. This is the first report of an investigation on speech discrimination in dyslexic adults with source estimates in the individual brain space. We found MMFs and late MMFs to all three speech-sound changes, generated in bilateral auditory cortices. Furthermore, across groups these MMFs and late MMFs were left-lateralized to the vowel deviant and the late MMF to the duration deviant. Contrary to our expectations, MMFs and late MMFs did not differ in source amplitudes or latencies between dyslexic and typical readers. However, the MMFs were associated with skills pertinent for reading. Stronger MMF source strengths in the left hemisphere were associated with better working memory skills across both groups and in the control group. This correlation seemed to originate specifically from the MMF to the duration deviant across groups, and from the verbal working memory component. In the dyslexic group, stronger MMF and late MMF in the right hemisphere correlated with better technical reading skills. To summarize, unlike several previous studies, we were not able to find evidence for poor phonological representations in dyslexia in the current sample. However, brain activity reflecting phoneme processing was associated with skills needed for fluent reading. This highlights the functional role of speech-related brain activity in reading and its impairments and promotes the utilization of the auditory MMF as a potential neural marker of abnormal reading.

### 4.1 Neural sources of MMF and late MMF

The sources of the MMFs indicated two response components: The MMF peaking at around 125–180 ms after change onset with a clear and narrow peak, the latency lying well within the established time range of the expected MMN/MMF (Kujala, Tervaniemi, & Schröger, 2007), having been associated with speech-sound discrimination (Kujala & Näätänen, 2010), and an additional broader, smaller response peaking at around 375–420 ms (termed late MMF). The late MMN, usually found in children’s responses (Cheour, Korpilahti, Martynova, & Lang, 2001; Volkmer & Schulte-Körne, 2018), has been reported only by few studies in adults at around 340–600 ms (Hill, McArthur, & Bishop, 2004; Hommet et al., 2009; Korpilahti, Lang, & Aaltonen, 1995; Schulte-Körne et al., 2001; Zachau et al., 2005). In adults, its functional role is still poorly understood. It was proposed to reflect linguistic processes as it was found to vowel changes, but not to tone changes (Hill et al., 2004; Korpilahti et al., 1995), although some studies have reported late MMNs to simple and complex tone changes (Schulte-Körne et al., 2001; Zachau et al., 2005). Due to the limited stimulus set of the current study that was not designed to investigate the role of the late MMF, the nature of this response remains to be investigated in more detail by future studies.

Our source estimates of both MMF and late MMF were located in primary and secondary auditory cortices (peak MNI coordinates corresponded to BAs 41, 21, 22) for all speech-sound changes. This is consistent with findings on MMN sources (Alho, 1995; Escera, Alho, Schröger, & Winkler, 2000). Our source modeling with distributed dipoles (MNE) confirms previous MMN/MMF findings from single-dipole models with a better spatial accuracy.

Our MMF lateralization analyses revealed stronger MMFs in the left than right hemisphere to vowel deviants and late MMFs to duration and vowel deviants in both groups, in line with previous studies suggesting left-hemispheric lateralization of speech-elicited MMFs (e.g., Näätänen et al., 1997; Shtyrov et al., 2000). Vowel duration changes make phonological distinctions in Finnish language (Kirmse et al., 2008; Näätänen et al., 1997), which explains the dominant use of the left hemisphere also for the duration deviants.

Furthermore, the LIs demonstrated left-dominant frequency discrimination in controls only. This is consistent with an earlier finding of left-dominant processing of frequency in speech stimuli (Sorokin, Alku, & Kujala, 2010). These results are compatible with the suggestion that frequency discrimination is linguistically relevant in Finnish language (Järvikivi, Vainio, & Aalto, 2010).

### 4.2 Group comparisons of sources

No group differences were found for the two responses of the MMF source amplitudes or latencies to vowel, vowel duration, or syllable frequency changes, suggesting no deficits in phonological representations in dyslexia, as opposed to the phonological deficit theory (Bradley & Bryant, 1983; Ramus, 2001; Ramus et al., 2013; Snowling, 2000). These results also contradict with many previous studies, which have shown diminished MMNs to speech-sound changes in dyslexia or dyslexia risk (Schulte-Körne & Bruder, 2010). Like our present study, the only one previous study comparing MMFs in dyslexic and control participants to speech-sound changes (/*ba*/ vs. /*da*/) also failed to find group differences (Paul et al., 2006). The authors suggested that this could have resulted from a too large stimulus difference that was too easy to discriminate for their dyslexic children. This is consistent with previous observations showing diminished MMNs in dyslexia for small but not for large stimulus differences (e.g., Baldeweg, Richardson, Watkins, Foale, & Gruzelier, 1999). The same could be the core reason for insignificant MMF source strength differences between groups in the present study, too. Alternatively or additionally, the dyslexic subsamples of the different studies might differ in terms of their phonological deficits, since not all dyslexics have such deficits (e.g., only one third of children with “pure” dyslexia had deficits in phonological representations in Ramus et al., 2013). Future studies should determine the prevalence of core phonological deficits vs. dysfunctions in accessing or associating phonemes during reading in dyslexia in large participant samples. For instance, it was shown that dyslexics who displayed normal-like MMNs to phoneme changes presented with meaningless visual stimuli had diminished responses to the same changes when they were accompanied with written input (Mittag, Thesleff, Laasonen, & Kujala, 2013). Therefore, it can be predicted that a larger proportion of dyslexics suffer from impairments in integrating and accessing of phonological information than merely from their poor representations.

In our previous study (Thiede et al., 2019) utilizing identical stimuli and paradigm as the current one, we found absent and atypical MMNs in infants at risk for dyslexia. This is quite a robust finding, since only ≈40– 70% of children at risk of dyslexia become reading impaired (DeFries & Alarcón, 1996). As we did not see such deficits in adult dyslexics, this suggests that neurobiological abnormalities in dyslexia may be more disruptive in infancy/childhood than in adulthood (see also, e.g., Lovio et al., 2010, reporting diminished MMNs to a range of speech stimuli in at-risk children). Possibly, speech development is originally delayed in dyslexia but speech processes become more normal by adulthood (Galaburda, LoTurco, Ramus, Fitch, & Rosen, 2006).

We expected to find group differences in MMF lateralization based on some previous studies on dyslexia (e.g., Kujala et al., 2003; Renvall & Hari, 2003; Sebastian & Yasin, 2008) or poor reading outcome (Maurer et al., 2009), but found no such effects. The results on laterality effects in dyslexia and related language impairments are mixed (Schulte-Körne et al., 2001; Sebastian & Yasin, 2008; Wilson & Bishop, 2018), which calls for larger sample sizes and careful phenotyping.

### 4.3 Correlation of source strengths with reading-related skills

We also aimed to determine whether reading-related measures are associated with MMF source strengths. With moderate sample sizes and a procedure addressing the correction of multiple comparisons, we found that larger MMF source amplitudes in the left hemisphere were associated with better working memory skills across both groups, suggesting the relevance of efficient working memory for speech processing. *Post-hoc* analyses showed that the association was mainly driven by the MMF to the duration deviant and the verbal component of working memory. Studies associating verbal working memory and MMN are rare and yielded mixed results. A diminished MMN was found to predict a working memory impairment (Ahveninen et al., 1999; Javitt, Doneshka, Grochowski, & Ritter, 1995). In addition, a larger MMN was found to be associated with increased verbal working memory performance in children (Watson, Titterington, Henry, & Toner, 2007). Yet, another study found no connection between the MMN and working memory (Light, Swerdlow, & Braff, 2007). These studies together with the present results call for further investigations on the complex interplay between speech processing, working memory, and reading.

For technical reading, we found associations only in the dyslexic group: Larger right-hemispheric MMFs and late MMFs correlated with better skills. However, it is notable that lack of this association in the control group could result from the lack of variation in the technical reading scores due to a ceiling effect (see Figure S 1). Previously, increased left-hemispheric MMN was associated with better word reading skills in children (Maurer et al., 2009), and longer MMN latency was associated with more errors in word and nonword reading across dyslexic and typical readers and within the dyslexic group (Baldeweg et al., 1999). The correlation between stronger right-hemispheric MMF sources and reading skills in our dyslexic group might suggest that some dyslexics have developed a right-hemispheric compensatory mechanism for speech processing that is also beneficial for their reading skills (Eden et al., 2004).

### 4.4 Limitations

The following limitations of the present study should be considered. The first one relates to the selection of the study sample. The participants of this study were adults between 18 and 45 years of age. Even though they were screened carefully for dyslexia to ensure that reading problems persisted into adulthood, many dyslexic adults may have found coping strategies for these problems since school age. Furthermore, our selected sample differed in IQ between the dyslexic and the typical-reading group. We addressed this by repeating our analysis with a performance-IQ -matched subsample and obtained similar results. Group sizes in this study were moderate consisting of 20+ participants in each group. Larger-scale studies should be aimed for, as the neuroimaging field suffers from replication failures of previous results obtained with small sample sizes (Kellmeyer, 2017). As the expected effect sizes are generally small, many of these studies might be underpowered. Second, the stimuli chosen for the current study have limitations, as discussed above. Stimuli that may be more sensitive to tap into the phonological impairment thought to be the predominant cause for dyslexia should be studied in the future. Lastly, future studies are needed to determine the nature of the late MMN/MMF.

### 4.5 Conclusions

To summarize, our results advanced with source-localization constraints from individual anatomical brain images support the suggestion of bilateral sources of the MMF to speech-sound changes in auditory cortices, as well as left-hemispheric lateralization of the MMF to vowel and vowel duration changes. We found comparable MMF strengths, latencies, and lateralization in typical and dyslexic readers, not confirming abnormalities in neural speech-sound discrimination in dyslexia, possibly due to stimuli not sensitive enough to probe these deficiencies, or a participant subsample not predominantly having phonological problems. However, we found correlations between the MMFs to speech-sound changes and reading-related skills, highlighting the connection of neural low-level speech processing and reading in adults, and promoting the use of MMFs in investigating reading-related brain processes.

## Data Availability

The code used for the analysis of this dataset is available.

https://github.com/athiede13/neural_sources

## CRediT author statement

**Thiede, A**.: Conceptualization, Investigation, Formal analysis, Data Curation, Writing - Original Draft, Visualization, Project administration, Funding acquisition. **Parkkonen, L**.: Resources, Formal analysis, Writing - Review & Editing. **Virtala, P**.: Conceptualization, Writing - Review & Editing, Supervision, Project administration, Funding acquisition. **Laasonen, M**.: Conceptualization, Writing - Review & Editing. **Mäkelä, J.P**.: Resources, Writing - Review & Editing. **Kujala, T**.: Conceptualization, Writing - Review & Editing, Supervision, Funding acquisition.

## Acknowledgements

The authors thank all participants that were involved in this study as well as all research assistants for their help with data acquisition and psychological testing, Marita Kattelus for her help with MRI acquisition, and the physician for examining the MRIs for incidental findings. The project was funded by Jane and Aatos Erkko foundation, Academy of Finland [project numbers 2764141 and 316970], KELA (The Finnish Social Insurance Institution), and University of Helsinki MRI measurement support. The first author was supported by the University of Helsinki Research Foundation and Arvo and Lea Ylppö foundation.

